# Photoacoustic 3D imaging detects potential microvascular injuries

**DOI:** 10.1101/2020.11.02.20224204

**Authors:** Yoshihiro Ishida, Yoshiaki Matsumoto, Yasufumi Asao, Aya Yoshikawa, Masako Kataoka, Takayuki Yagi, Kenji Kabashima, Masakazu Toi

## Abstract

The photoacoustic imaging (PAI) system is an emerging imaging modality that can be useful in clinical diagnostic testing. Previous studies have shown utility of PAI in diagnosing breast cancer, visualizing vascular change in ageing, and planning flap reconstruction surgeries. In this study, we show that PAI can be used to non-invasively visualize microvascular injuries in smokers, and in a vasculitis patient.

We used two prototype PAI systems, PAI-03, and PAI-04 in this study. These systems were equipped with hemisphere-detector arrays, and used lasers at wavelengths of 795 nm (PAI-03), 756 and 797nm (PAI-04). Healthy volunteers and a patient diagnosed with vasculitis were enrolled upon obtaining written informed consent. The whole hand of the volunteers was scanned.

We noted there were unique circular structures that we coined “minute signals”. There were higher number of minute signals in smokers than in non-smokers. Enhanced minute signals were noted in the patient with vasculitis. In this patient, minute signals were suggested to have lower oxygenated hemoglobin content, likely suggesting blood clots or hemorrhage. Our study shows PAI systems can visualize unappreciated abnormalities in fine vasculatures non-invasively. Use of PAI can extend to other clinical entities with suspected vascular involvement.

## Introduction

The photoacoustic imaging (PAI) system is a diagnostic imaging system that can produce high resolution images of the subcutaneous vasculature^1-8)^. Unlike conventional computed tomography (CT) or magnetic resonance imaging (MRI) systems, the PAI systems do not use contrast agents and can be used to create vasculature images non-invasively. Also, unlike ultrasound doppler method, it does not measure blood flow but rather detect hemoglobin distribution. It can visualize not only blood vessels, but also stationary absorbers such as hematomas by visualizing the local distribution of hemoglobin. In order to obtain good three-dimensional (3D) images with photoacoustic technology, the photoacoustic waves generated by the light irradiation should be received from the whole 3D directions^9)^. We have reported the effectiveness of hemispherical detector arrays (HDA) that can surround the subject and receive the photoacoustic wave generated from the subject^3-8)^. There are reports that this device can be used to plan the flap reconstruction procedure^4)^, and to explore the anastomosing vessels for lymphaticovenular anastomosis (LVA)^5)^. Exploratory studies used PAI to visualize angiogenesis induced by breast cancer^6)^ and show the relationship between palmar artery and age^7)^. These studies have shown that PAIs can analyze detailed peripheral vascular network structures.

Visible nodules^10)^ in the fingers have been reported in COVID-19 cases resulting from thrombosis of peripheral vessels. Microvascular disorders are implicated in acute diseases such as disseminated intervascular coagulation. In addition, chronic diseases such as diabetes and smoking are known to cause microvascular disease. However, there are only indirect indicators of microvascular injuries, such as blood tests. For a more detailed diagnosis, high-definition imaging modality will play an important role. We have reported that the PAI can visualize vessels in the palm and analyze the common and proper palmar digital arteries after removing subcutaneous vein networks^11)^ within the three-dimensional (3D) images. In the present study, we focused on subcutaneous veins and found “knot” signals in the vicinity of the veins. We present two separate clinical studies (1. in healthy subjects and 2. in patients with vasculitis) in which knots were detected. We propose that these knots are indicative of microvascular injury, and that PAIs can be used to diagnose the undetected microvascular injuries.

## Material and Method

### Device Configuration

We used two PAI systems with hemispherical detector arrays (HDA), PAI-03^7)^ and PAI-04^6)^, whose configurations were previously described. As reported, when the horizontal plane is the xy plane and the vertical direction is the z axis, the laser light is irradiated along the Z axis from the bottom of the HDA. In PAI-03, an Alexandrite laser with a wavelength of 795 nm was used for the measurement. In PAI-04, we used two Ti:Sa lasers optically pumped using a Q-switched Nd:YAG laser, which can select wavelengths from 700 to 900 nm. We selected two wavelengths, 756 and 797 nm, to construct an improved hemoglobin saturation distribution image. While 756 nm is the local maximum of the absorption coefficient of deoxidized hemoglobin, 797 nm is the isosbestic point of oxyhemoglobin and deoxidized hemoglobin.

### Body position while scanning

The body position while scanning was the same as previously reported. The palm was placed on a holding cup to keep the subjects’ palm parallel to the xy-plane. The imaging area was within a circle of 140 mm in diameter for both devices. The scanning time was approximately 2 minutes. The subject’s hand was fixed in a holding cup and either right or left hand was scanned. Researchers decided which hand to be scanned at researchers’ discretion.

### Imaging method

Universal back projection^12)^ (UBP) was used to reconstruct the 3D PA image obtained in the palm. The voxel size at UBP reconstruction was 0.125 mm for both devices. For the reconstructed voxel data, the maximum intensity projection method (MIP) which projects the maximum voxel value along the z-axis direction onto the xy plane was applied. Since PAI-03 is an evaluation of a single wavelength, we created a two-dimensional image in black and white grayscale display in which the voxel values corresponded to the image brightness values. In PAI-04, the image of weighted S-factor defined in the previous paper^13)^ was used. Since the optical fluence in vivo was not evaluated, and the absolute value of oxygen saturation could not be calculated. Therefore, we only evaluated the relative magnitude of oxygen saturation in the region of interest (ROI).

### Ethics

This study was approved by the Ethics Committee of the Kyoto University Graduate School of Medicine (UMIN000018893 and UMIN000022767), and we obtained written informed consent from all subjects. This study was conducted in accordance with the Declaration of Helsinki. The subjects of the UMIN000018893 study were healthy volunteers whose palms were imaged using the PAI-03 system. The subjects of the UMIN000022767 study were both healthy volunteers and a patient with skin disease whose palms were imaged using the PAI-04 system.

The study in UMIN000018893 is referred to as CS1 (Clinical study-1) and the study in UMIN000022767 as CS2 (Clinical study-2) in this report.

### Subjects

#### [CS1]

PAI-03 was used in this experiment. We recruited healthy men and women of age from 20s to 50s. We recruited subjects in blocks so that the number of men and women included for each age group would be 2-3 in each block. Health status of the subjects were self-reported.

Nineteen persons (8 men and 11 women) were recruited. Mean age of men was 36.1 years old (range: 22-56) and mean age of women was 38.5 years old (range: 25-59). Two persons were smokers and the other 17 were non-smokers.

#### [CS2]

PAI-04 was used in this experiment. 2 healthy females (age: 30s and 40s, respectively) were included as control. A patient with vasculitis (age: 60s) affecting the lower limbs was recruited.

### Data analysis

Quantitative analysis was performed on [CS1] data, and [CS2] was assessed only qualitatively given the small number of subjects enrolled in [CS2].

Grayscale MIP images from [CS1] were manually inspected for the knots. We defined “knots” using two separate criteria and analyzed them independently; (See Supplementary figure 1.)

**Figure 1.**
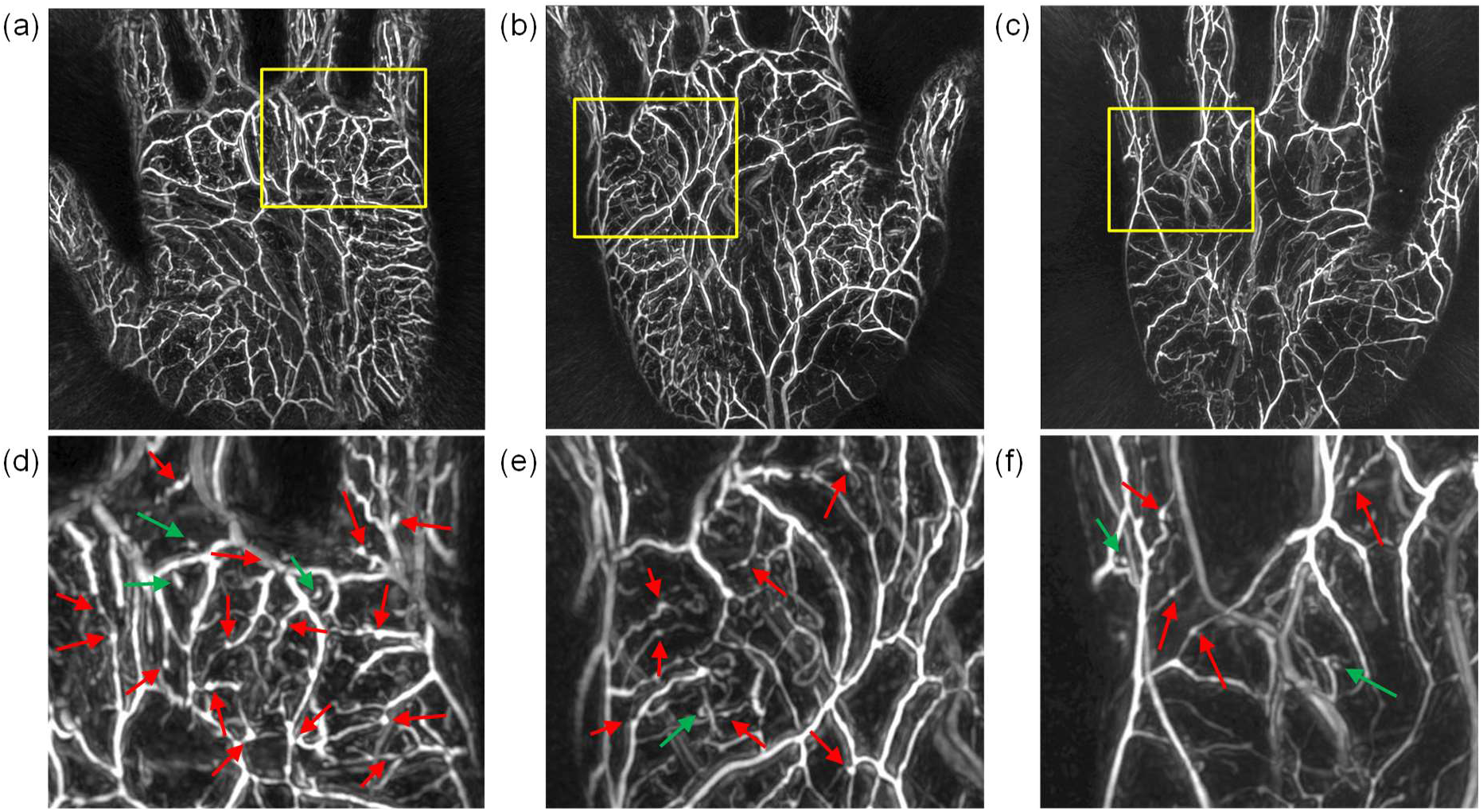
Palm vasculature obtained from young subjects. (a) 20s male, smoker, (b) 30s male, non-smoker, (c) 30s female, non-smoker. (d), (e) and (f) are enlarged views of the parts surrounded by the yellow squares of (a), (b) and (c). In these (d), (e) and (f), the red arrows are the knots extracted by the criterion 1, and the green arrows are the knots extracted by the criterion 2.

[Criterion 1] Circular structures along the blood vessels were defined as “knots”. However, if the structures were located on the branching points of the blood vessels, they were not counted as knots because they may reflect normal anatomical structure.

[Criterion 2] Circular structures not in contact with a blood vessel were defined as “knots”. Circular structures in contact with a vessel were also included if the center line of the vessel and the center of the circle are far apart.

The knots were counted based on the above criteria, and the results were compared with respect to smoking status or gender difference. Numeric data was analyzed by Mann-Whitney U test with P value < 0.05 considered significant.

## Results

### Results of CS1 (Healthy Subjects)

Nineteen palms were scanned (See Supplementary figure 2). There are more knots in smokers than non-smokers and males than females (Fig 1).

**Figure 2.**
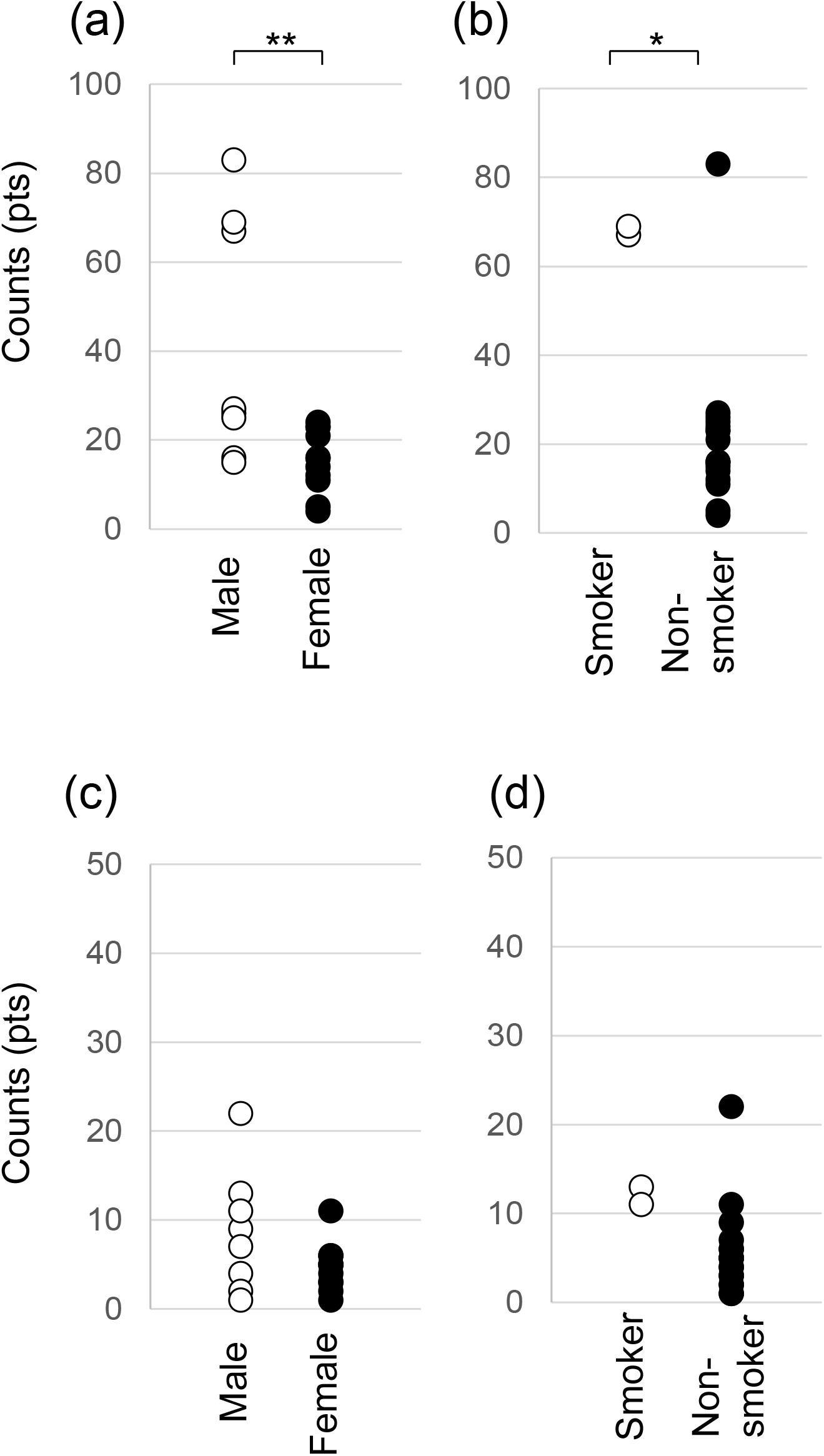
Quantitative analysis of the knots. (a) Comparison between male and female by criterion 1. A significant difference was observed (p<0.01). (b) Comparison of smoking status by criterion 1. A significant difference was observed (p<0.05). (c) Comparison between male and female by criterion 2. No significant difference was obtained. (d) Comparison of smoking status by criterion 2. No significant difference was obtained.

Next, we analyzed the relationships between the number of knots and clinical variables. Figures 2 (a)-(d) compare smoking statuses and gender differences using criteria 1 and 2, respectively. knots defined by criterion-1 showed fewer in females than in males (p=0.00632) and in non-smoker than in smoker (p=0.04582). It was not correlated with age (p =0.708 in male and p=0.0797 in female by Pearson’s test). (See Supplementary figure 3 (a).)

**Figure 3.**
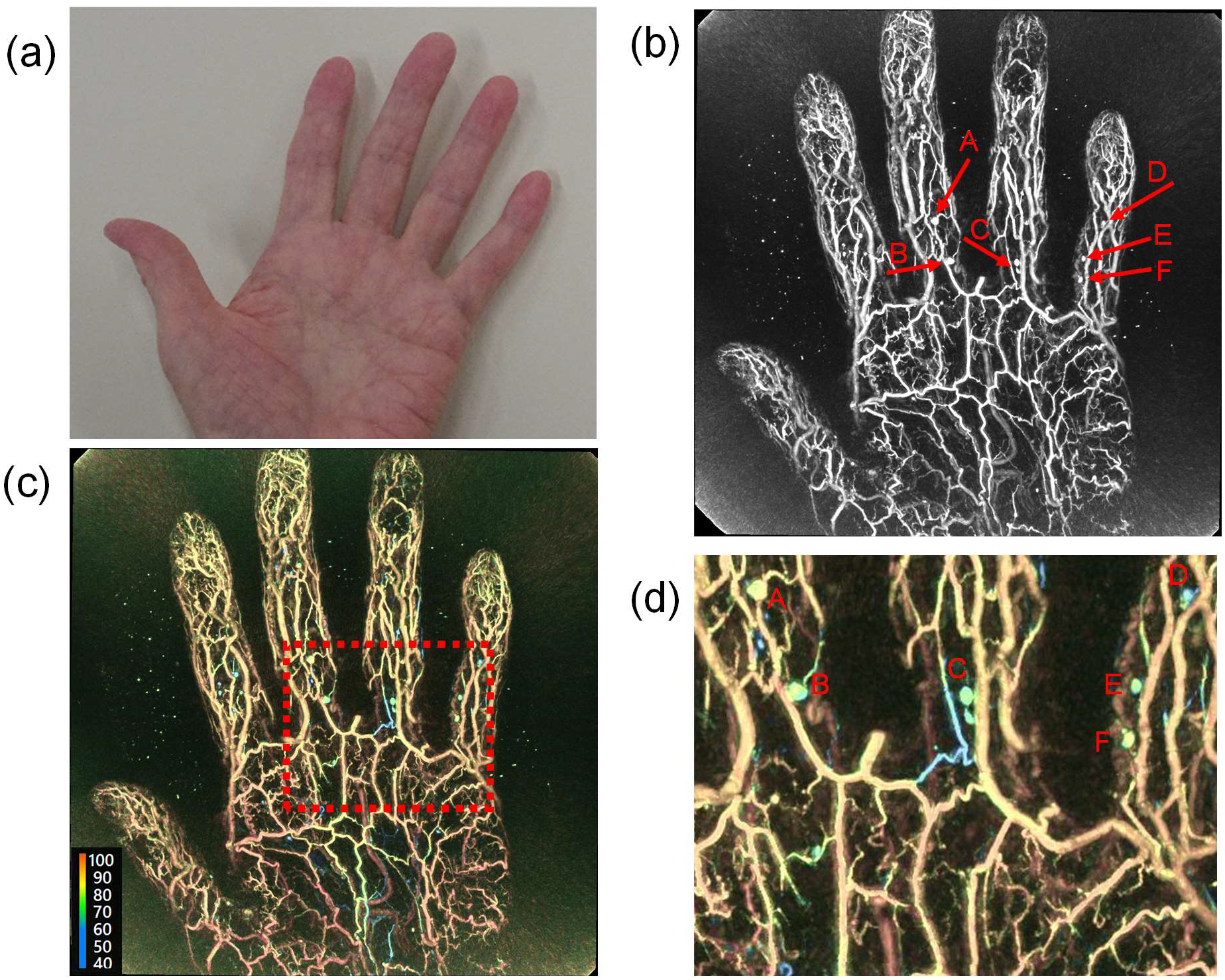
Images of a patient showing vasculitis symptoms on his foot but not in the hand. (a) A photograph of the palm. No visible symptoms were observed on the palm. (b) MIP image of the hand (wavelength: 797nm) Red arrows show typical knots. (c) Oxygen-saturation of the same image (Red: high oxygen content, blue: low oxygen content) (d) An enlarged view of (c)

In contrast, the number of knots defined by criterion 2 did not show difference between males and females (P=0.2135), or smoker and non-smoker (P=0.05303). It was also not correlated with age (p =0.546 in male and p=0.463 in female by Pearson’s test). (See Supplementary figure 3 (b).)

### Results of CS2 (Vasculitis Patient)

First, we qualitatively compared the images acquired from healthy subjects by PAI-03 and PAI-04. On both platforms, the female subjects showed fewer knots and the blood vessels were smoother (supplementary figures 4 and 5). We concluded that PAI-04 can capture blood vessel images with the same imaging performance as PAI-03.

Then, we assessed images captured from the palm of a vasculitis patient, who showed clinical symptoms on the foot but not in the hand (Figure 3 (a) and (b)). PAI-04 is capable of inferring oxygenation status of hemoglobin by using two wavelengths; these knots show relatively lower oxygenation status (Figure 3 (c) and (d)).

## Discussion

It has been widely reported that PAI can be used to image subcutaneous microvasculature. In this study, we focused on the knots around the vasculature and analyzed them both quantitatively and qualitatively.

The limitation of this study was that we could not analyze knots histologically. However, we can safely assume that knots were from red blood cells or hemoglobin clustered in a circle, given that the knots were observed at a depth and position compatible with known anatomy of the human hand. These knots may reflect the morphology of the venous valves^14)^, or they may reflect abnormal blood flow in the microcirculatory disturbances that occur in the peripheral veins and venules.

In the CS1 (healthy subjects), analysis with criterion 1 suggested that the number of knots between men and women and between smokers and nonsmokers were significantly different. The low knot count in women may be attributable to female sex hormones’s effect on blood vessels^15)^. As for the smokers, this may be due to the microvascular injuries, i.e., a thrombus-like retention of blood caused by smoking. Smoking has been reported to increase blood pressure and heart rate but reduce skin surface temperature and subcutaneous blood flow^16)^. It can be assumed that this places considerable strain on the small blood vessels under the skin, causing impaired blood flow. As a result, venous blood may flow retrograde and the venous valves may derive distention. In addition, it has been reported that vascular endothelial cells are weakened in smokers and patients with vasculitis^17-18)^. This effect may lead to micro-thrombus-like retention of blood. These differences in vascular morphology could be detected by PAI.

On the other hand, the isolated knots shown in criterion 2 were not significantly different regardless of the subject. The reason for the isolated knot may be due to the limitations of the device’s performance, which prevented us from imaging fine veins with diameters close to the capillaries, and only the enlarged knot area was imaged. Although the number of microthrombi occurring in these small venules may not be lifestyle-dependent, further investigation should be needed.

In the CS2 patient test, the reason for the clearer knot in the patient than in the healthy person may be that the patient had more advanced microcirculatory injuries. It is conceivable that blood vessels are weakened by vasculitis, and microcirculation disorders are likely to occur. In addition, since this circulatory disorder has a lower oxygen saturation than that of nearby veins, these knots may reflect subclinical thrombi or hemorrhage.

As there are no other modalities that allow us to observe microblood flow disturbances in peripheral veins, we must establish the evidence for this study by increasing the number of cases in the future. Another limitation of this paper is that we were unable to assess differences in age group and race among subjects over 75 years of age, who are generally classified as elderly. The further studies are needed to expand the scope of the present study and increase the number of subjects. We speculate that PAI may be useful in early diagnosis of COVID-19 that is reported to cause dysfunction of endothelial cells^19)^.

In summary, we used a PAI equipped with a hemispherical detector array to analyze palmar vascular images. Knots were found in the vicinity of the subcutaneous veins, which may have captured previously undetectable microcirculatory disturbances. These knots were suggested to be different between men and women and between smokers and non-smokers. Distinct bums were observed in asymptomatic patients with vasculitis. The oxygen saturation results suggest that the knots are thrombi or hemorrhage. Since PAIs can diagnose vascular disorders noninvasively, use of PAIs can be extended to early diagnosis of various diseases.

## Data Availability

Relevant data are provided upon reasonable request.

## Acknowledgements

This work was partially funded by the ImPACT Program of the Council for Science, Technology and Innovation (Cabinet Office, Government of Japan). The authors thank T. Kosaka and K. Kobayashi for coordinating the activities for this clinical research. and Canon, Inc., for providing the PAI-03 and PAI-04 systems.

---

Supplementary figure 1

Schematic diagram representing criterion 1 and criterion 2. In criterion 1, a circular signal was counted when there was a circular signal directly above a blood vessel. For criterion 2, a circular signal was counted when a circular signal existed at a position apart from or off the center of a blood vessel.

Supplementary figure 2

Compilation of all images taken in CS1, where M indicates male and F indicates female. The numbers represent the age group.

Supplementary figure 3

The count results of the knots as a function of the subject’s age. (a) is derived from the result of [Criterion-1], and (b) is that of [Criterion-2]. The smokers’ results were indicated as an asterisk “*” in the upper right of the plot. Open circles indicated for males and closed circles for females.

Supplementary figure 4

The 1^st^ control image of CS2 obtained by PAI-04. (40s, female)

Supplementary figure 5

The 2^nd^ control image of CS2 obtained by PAI-04. (30s, female)

