## Supplementary Figures for "Photoacoustic 3D imaging detects potential microvascular injuries"

Supplementary figure 1

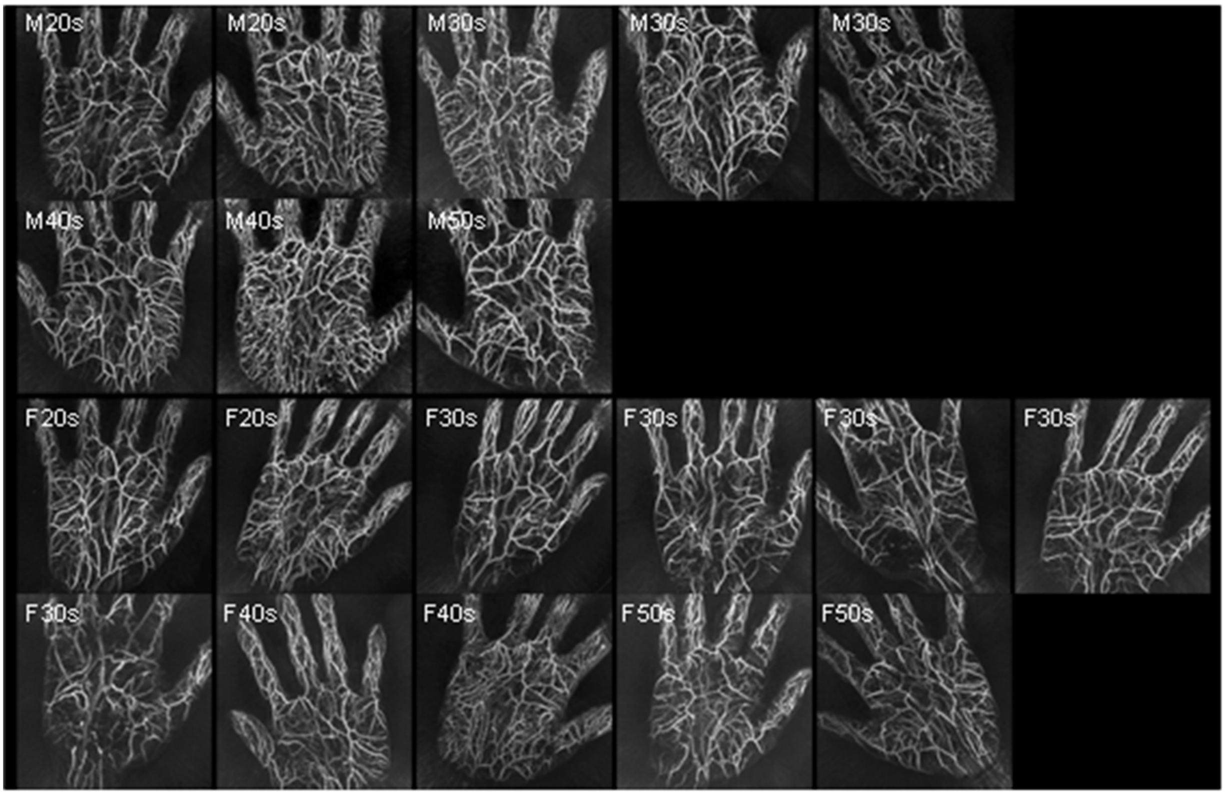

Supplementary figure 2

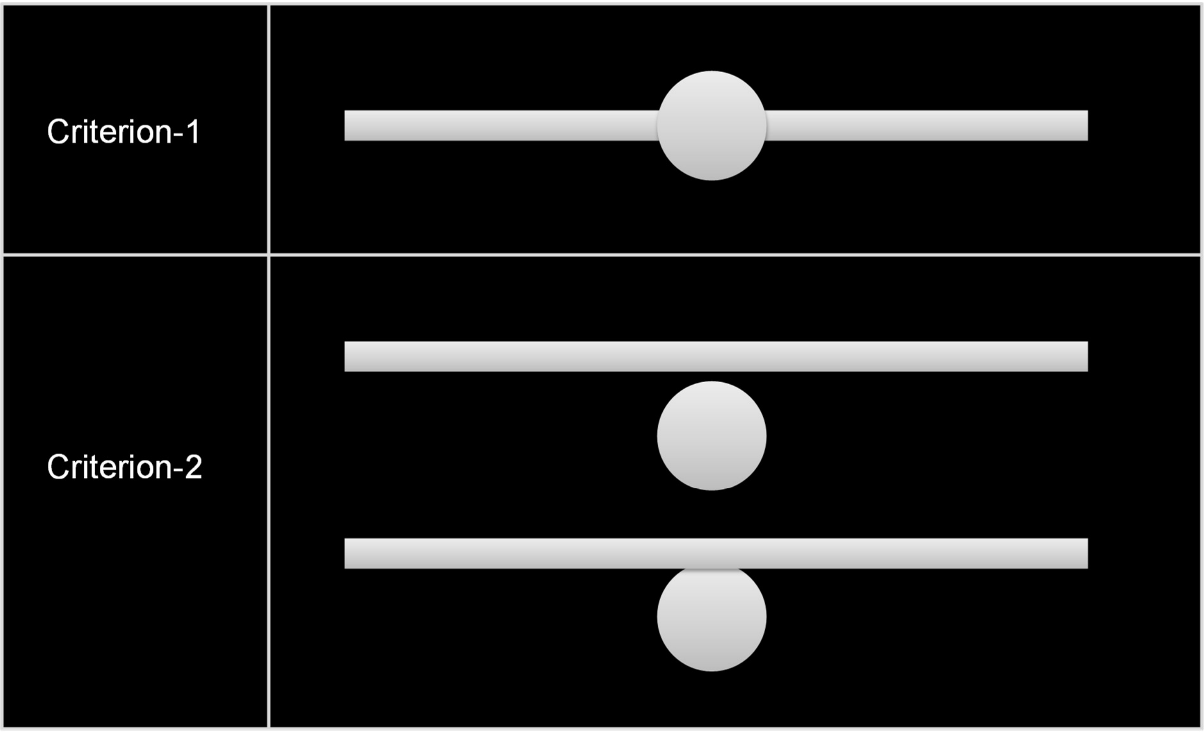

Supplementary figure 3

(a)

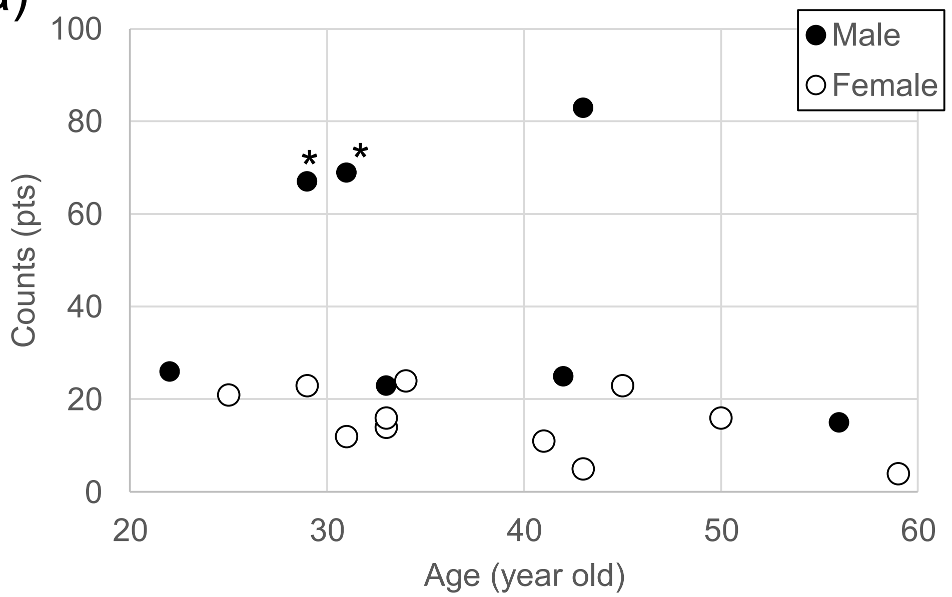


(b)

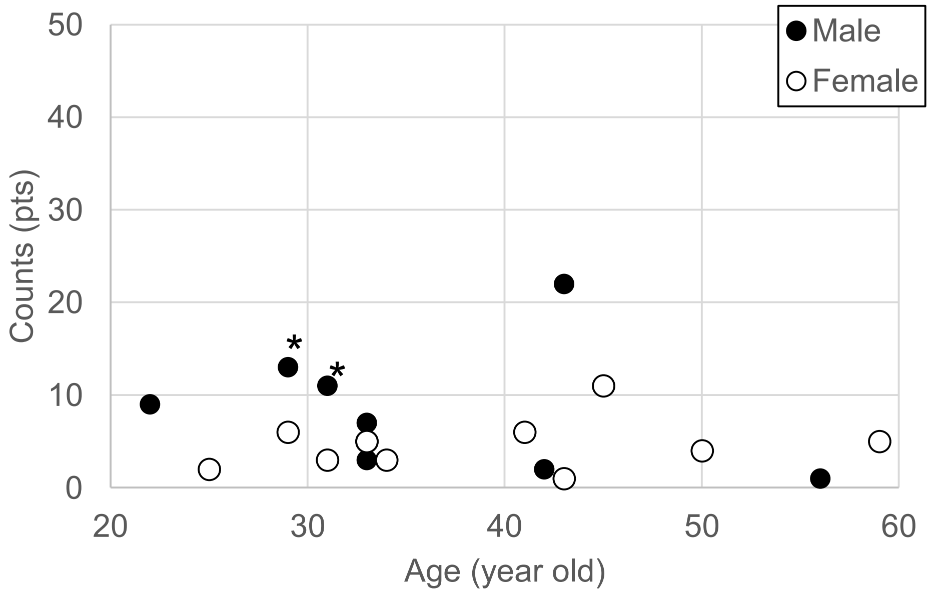

Supplementary figure 4

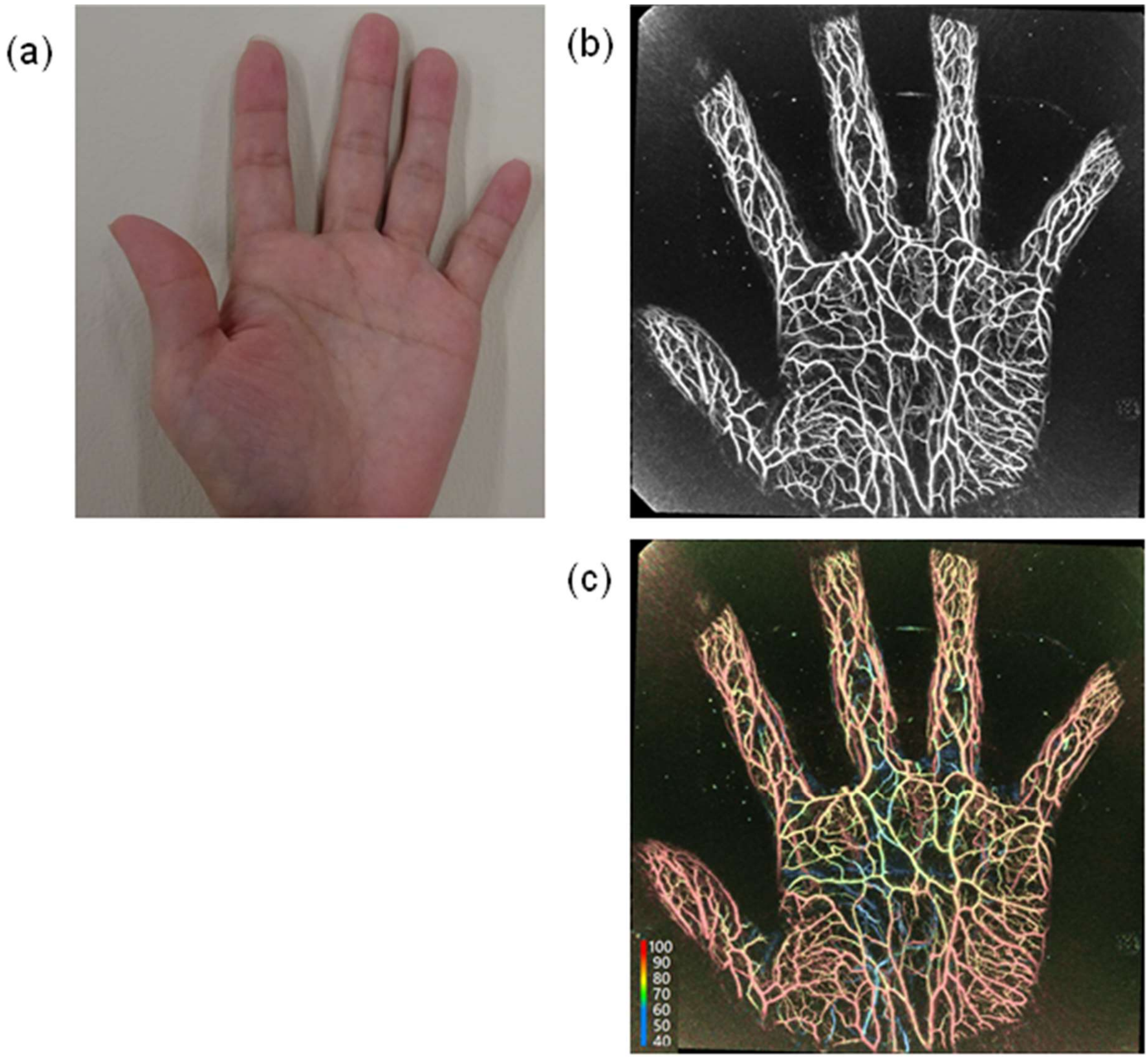

Supplementary figure 5

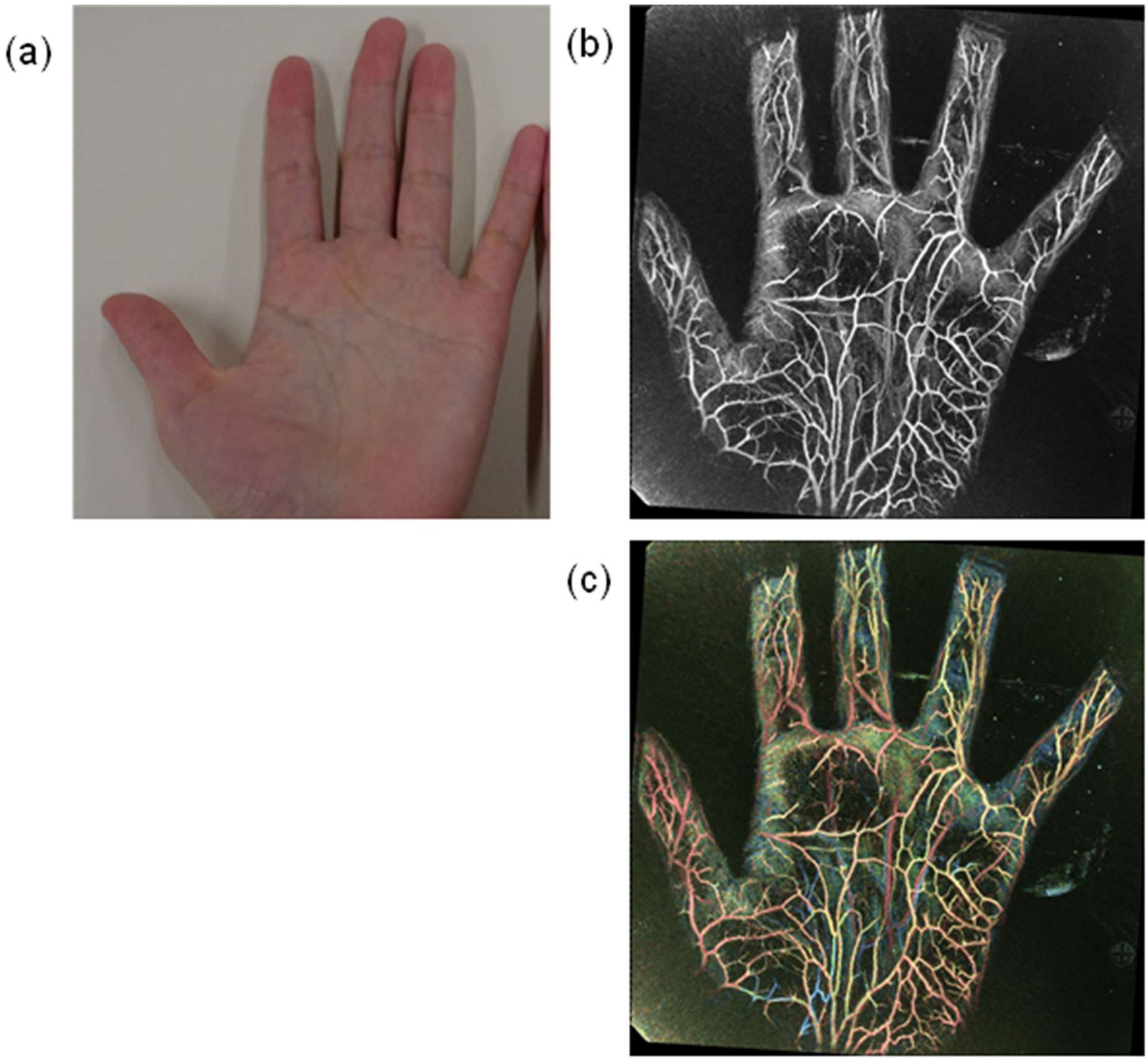
